# 11β-HSD1 inhibitor efficacy in type 2 diabetes is cortisol-dependent

**DOI:** 10.1101/2024.05.10.24307180

**Authors:** Atinuke Wilton-Waddell, Layal Abi Farraj, Elton JR Vasconcelos, Emily Byrne, Angela E Taylor, Adrian Freeman, Damla Etal, Paul M Stewart, Wiebke Arlt, Ramzi Ajjan, Ana Tiganescu

**Affiliations:** Leeds Institute of Cardiovascular and Metabolic Medicine, University of Leeds, Leeds, UK; Department of Pathology and Laboratory Medicine, University of California Los Angeles, USA, Leeds Institute of Medical Research, University of Leeds, Leeds, UK; Leeds Omics, University of Leeds, Leeds, UK; Institute of Metabolism and Systems Research, University of Birmingham, Birmingham, UK; Emerging Innovations Unit, Discovery Sciences, BioPharmaceuticals R&D, AstraZeneca, Cambridge, UK; Translational Genomics, Centre for Genomics Research, Discovery Sciences, BioPharmaceuticals R&D, AstraZeneca, Gothenburg, Sweden; NIHR Biomedical Research Centre, Leeds Teaching Hospitals NHS Trust, Leeds, UK; Faculty of Medicine and Health, University of Leeds, Leeds, UK; IMRC Laboratory of Medical Sciences, London, UK; Institute of Clinical Sciences, Faculty of Medicine, Imperial College London, London, UK

## Abstract

Cortisol excess drives multiple adverse effects including hypertension, dyslipidemia, and delayed wound healing. Activation of cortisol by the enzyme 11β-hydroxysteroid dehydrogenase type 1 (11β-HSD1) has shown promise as a therapeutic target for these comorbidities but clinical progress has been hampered by variable 11β-HSD1 inhibitor efficacy. Here, transcriptomic profiling of 11β-HSD1 target genes in primary skin fibroblasts as well as skin biopsies from type 2 diabetes individuals treated with the selective 11β-HSD1 inhibitor AZD4017 provide detailed mechanistic insights highlighting new areas of therapeutic potential. We report correlations between changes in 11β-HSD1 target gene expression, blood pressure, lipids, and wound healing with 1) cortisol levels (serum cortisol / dehydroepiandrosterone sulfate) and 2) peripheral 11β-HSD1 activity (serum cortisol / cortisone). Finally, we demonstrate that baseline cortisol levels and changes in placebo group cortisol levels are key determinants of 11β-HSD1 inhibitor efficacy. In conclusion, our findings pave the way for more effective targeting of 11β-HSD1 inhibitor treatment, improving the accuracy of future clinical studies. Larger trials of longer duration are now warranted to fully explore the therapeutic potential of 11β-HSD1 inhibitors across a range of cardiometabolic and age-associated indications.

## 1. INTRODUCTION

Glucocorticoid (GC) excess drives multiple systemic pathological processes including hyperglycaemia, hypertension, elevated intraocular pressure, hepatic steatosis, muscle atrophy, osteoporosis, delayed wound healing and skin atrophy[1]. These effects are mediated at the tissue level by 11β-HSD enzymes which mediate the conversion to active cortisol from cortisone (11β-HSD1), while also inactivating cortisol to cortisone (11β-HSD2) independently of circulating hormone levels[2]. Reducing GC exposure through 11β-HSD1 inhibition showed therapeutic promise across several cardiometabolic outcomes, including lowering weight, blood glucose, blood pressure and cholesterol[3–10], however, variable efficacy and inconsistent effect sizes have delayed clinical progress.

Our pre-clinical studies found that 11β-HSD1 inhibition improved wound healing during GC excess[11]. More recently, we reported improved wound healing, together with lower blood pressure and healthier lipid profile following 11β-HSD1 inhibition in people with type 2 diabetes[7]. However, mechanistic detail to better understand the genes and pathways regulated by 11β-HSD1 is lacking. Addressing this knowledge gap is an important step towards developing a new generation of therapeutics.

Transcriptome profiling is a powerful technique to understand human pathologies at a molecular level. To elucidate the mechanistic pathways regulated by 11β-HSD1 in skin, we used human skin fibroblasts integral to the wound healing process[12], and skin from people treated with AZD4017 (selective 11β-HSD1 inhibitor) for the first transcriptomic profiling of 11β-HSD1 targets. Furthermore, we compared changes in the 11β-HSD1 skin transcriptome, wound healing, blood pressure and lipids with changes in cortisol levels (serum cortisol/dehydroepiandrosterone sulfate; F/DHEAS)[13, 14] and peripheral cortisol activation by 11β-HSD1 (serum cortisol/cortisone; F/E)[15]. We aimed to test whether people with low cortisol levels (low F/DHEAS) would be less responsive to 11β-HSD1 inhibition, and whether changes in cortisol levels in the placebo group could affect 11β-HSD1 inhibitor efficacy. If so, F/DHEAS and F/E could be useful biomarkers to improve 11β-HSD1 inhibitor targeting and future clinical study robustness.

## 2. METHODS

### **2.1.** Study participants

The study was conducted according to the Declaration of Helsinki principles, approved by the North West Greater Manchester Central Research Ethics Committee (17/NW/0283), and following written informed consent. Participants were randomised (double-blind) to PCB (n=14) or 400 mg bi-daily AZD4017 (n=14) for 35 days. At day 0 (baseline, pre-treatment), two 3 mm diameter full-thickness lower outer arm punch biopsies were conducted under local anesthesia, repeated in the contralateral arm at day 28. Wounds were treated with a breathable dressing for 24 hours and imaged on day 2 post-wounding. One biopsy per participant was immediately snap frozen in liquid nitrogen for RNA-seq studies. Full study details and participant demographics as previously reported[7].

### **2.2.** Human dermal fibroblasts

All reagents were obtained from Sigma Aldritch (Gillingham, UK). Primary human dermal fibroblasts (HDF) were cultured from abdominal skin of Caucasian female donors (42.4 ± 5.7 years) obtained by Bradford Ethical Tissue (Yorkshire & The Humber - Leeds East Research Ethics Committee 07/H1306/98). HDF were grown at 37°C, 5% CO2, in high-glucose DMEM + 10% fetal calf serum + 1% Pen-Strep. At 80% confluence, cells were passaged 1:3 at as required.

#### 2.2.1. Gene expression

HDF (biological n=3) were treated for 96 hours with vehicle (0.4% ethanol), 10ng/ml IL-1β, IL-1β + 200nM cortisone, IL-1β + cortisone + 1µM selective 11β-HSD1 inhibitor AZD4017, IL-1β + 100nM cortisol, or AZD4017. IL-1β treatment was required to induce sufficient 11β- HSD1 activity as previously described[16] and reflective of the inflammatory phase of wound healing.

#### 2.2.2. RNA extraction and reverse transcription

Total RNA was extracted using Trizol in combination with the PureLink RNA Mini Kit, following manufacturer’s protocol. For qPCR, cDNA was prepared to a final concentration of 10ng/μl with a Tetro cDNA Synthesis Kit, following manufacturer’s protocol.

#### 2.2.3. qPCR

Samples were loaded in 10μl volumes; 1μl cDNA (1ng/μl final concentration) + 5μl 2X SensiFAST™ Probe Hi-ROX mix (Bioline, UK) + 0.5μl 11β-HSD1 (Hs01547870_m1) or 0.15μl 18SrRNA (Hs03003631_g1) TaqMan primer/probe in RNase free water.

qPCR was conducted using the ABI 7900 system (Perkin-Elmer, UK) with the cycle parameters: 5 min 95 °C, 40 cycles of 10 sec 95 °C then 50 sec 60 °C. Ct values were averaged (triplicate) and normalized to 18S rRNA (ΔCt).

#### 2.2.4. Fluidigm qPCR

Fluidigm qPCR was conducted for validation on 246 (of 611) differentially expressed RNA-seq genes and 32 control genes (Supplemental Table 1) from HDF treated with IL-1β + cortisone or IL-1β + cortisone + AZD4017 (biological n=3).

Pre-amplification was conducted in 5μl volumes; 1μl TaqMan PreAmp Master Mix (5X, Thermo Fisher, Loughborough, UK) + 1.25μl pooled TaqMan gene expression assay (0.2X final assay concentration) + 1.25μl cDNA (final concentration 2.5ng/μl) + 1.5μl distilled water (in triplicate). Cycle parameters: 2 min 95°C, 14 cycles of 15 sec 95°C then 4 min 60°C. Samples were diluted 1:5 with 20μl 1X TE buffer, following manufacturer’s protocol. Sample Mix and Assay Mix were prepared following manufacturer’s protocol and 5µl each was loaded onto chips and run on a BioMark qPCR reader.

Following standard curve verification, data were obtained as Ct values and ΔCt values calculated normalizing to 18S rRNA. For RNA-seq validation, log read count values were correlated against Fluidigm qPCR Ct values from the same RNA sample.

#### 2.2.5. 11β-HSD1 activity

HDF (biological n=10) were treated for 72 hours with 200 nM cortisone, cortisone + 10ng/ml IL-β or cortisone + IL-β + 1µM ADZ4017 with tracer amounts of [3H] cortisone (8000cpm, specific activity 74.0 Ci mmol–1; NEN, Boston, MA) in duplicate. Steroids were extracted from the medium with dichloromethane, separated by thin-layer chromatography (chloroform:ethanol ratio 92:8) and the % conversion cortisone to cortisol calculated by scintillation with a MicroBeta2 (Perkin Elmer, UK).

#### 2.2.6. RNA-seq

RNA-seq was conducted on HDF treated with vehicle, IL1-β, IL1-β + cortisone, or IL1-β + cortisone + AZD4017 for 96 hours (biological n=3). RNA quality was confirmed by Agilent TapeStation (RIN > 8) and paired-end cDNA libraries were generated with a TruSeq Stranded mRNA Library Prep Kit.

cDNA libraries were sequenced by Illumina HiSeq 3000 (6 samples per lane). FASTQ files were quality-checked using FastQC and aligned to the hg19 human reference genome using TopHat. Gene matrices were generated using Subread and differentially expressed genes were identified using pairwise EdgeR comparing vehicle vs. IL-β, IL-β vs. IL-β + cortisone, and IL-β + cortisone vs. IL-β + cortisone + ADZ4017.

Target pathways were identified using over-representation analysis with ConsensusPathDB software[17]. All 611 differentially expressed genes were included in the analysis.

### **2.3.** Human skin

#### ***2.3.1.*** RNA-seq

Human skin biopsies were processed for total RNA using the RNAdvance Tissue Kit (Beckman Coulter, High Wycombe, UK)) with quantification (>25ng/µl) and quality confirmed (DV200 >30%) using the RNA Analyser Kit (Agilent Technologies, Cheadle, UK). cDNA libraries were generated by reverse transcription using Kapa RNA HyperPrep Kit with RiboErase (Roche, Welwyn Garden City, UK) and the Tecan Fluent Liquid Handler System (Tecan, Reading, UK). Libraries were quantified using the Next Generation Sequencing Kit using a 96-channel Fragment Anaalyser (Agilent Technologies) and the Qubit DNA High Sensitivity Kit (Fisher Scientific, Loughborough, UK). Libraries were diluted to 1.8nM and sequenced by NovaSeq 6000 using S1 Reagent Kits and 100 cycles (Illumina, Cambridge, UK), with average reads per sample of 73.5 million.

FASTQ files were quality-checked using FastQC and aligned to the reference genome with Salmon. Differentially expressed genes were identified using pairwise DESeq2 comparing baseline (untreated, day 0) vs. treatment day 28 in participants assigned to the AZD4017 arm, and baseline vs. day 28 in participants assigned to the placebo arm. Target pathways were identified using over-representation analysis with ConsensusPathDB software. Due to the antiproliferative effects of GC on multiple skin cell types, differentially expressed genes included 25 histones, resulting in a bias towards cell cycle pathways. To enable the detection of other pathways, these histones were excluded from the main analysis(all remaining 814 differentially expressed genes were included).

To investigate the impact of cortisol levels and 11β-HSD1 activity on AZD4017 efficacy, three AZD4017-treated participants with low baseline cortisol levels (who could be less responsive to 11β-HSD1 inhibition) and two placebo-treated participants with a relatively large reduction in cortisol levels (as anticipated for the AZD4017 group) were excluded from the first analysis (Supplemental Figure 1). These five potential confounders were then reintroduced for a second analysis correlating the change in 11β-HSD1 target gene expression (log2 fold-change) with the change in cortisol level and 11β-HSD1 activity.

### **2.4.** Statistical methods

Grouped analyses were performed using 95% confidence intervals and a one-way analysis of variance mixed-effects model with *post hoc* testing corrected for multiple comparisons using Sidak’s test (GraphPad Prism, La Jolla, California). Correlations were analyzed using Pearson’s correlation testing (95% CI).

## 3. RESULTS

### 3.1. Human dermal fibroblasts

HDF are GC target cells and key regulators of skin homeostasis and wound repair. HDF studies were conducted for a more detailed mechanistic analysis in this important skin cell type, compared to subsequent *in situ* skin studies.

#### 3.1.1. Validation

11β-HSD1 mRNA was induced 3-fold by IL-1β, with a further 2.2-fold induction when treated concomitantly with cortisol or cortisone (Figure 1A), as previously described[16]. Consistently, 11β-HSD1 activity also increased by 4.6-fold with IL-1β, a further 2-fold with IL-1β + cortisol, and was fully inhibited by ADZ4017 (Figure 1B). Validation of 246 (of 611) RNA-seq and 32 control genes demonstrated a strong correlation between RNA-seq read counts and Ct values (qPCR) from the same sample (r = -0.79, p<0.001, Figure 1C).

**Figure 1.**
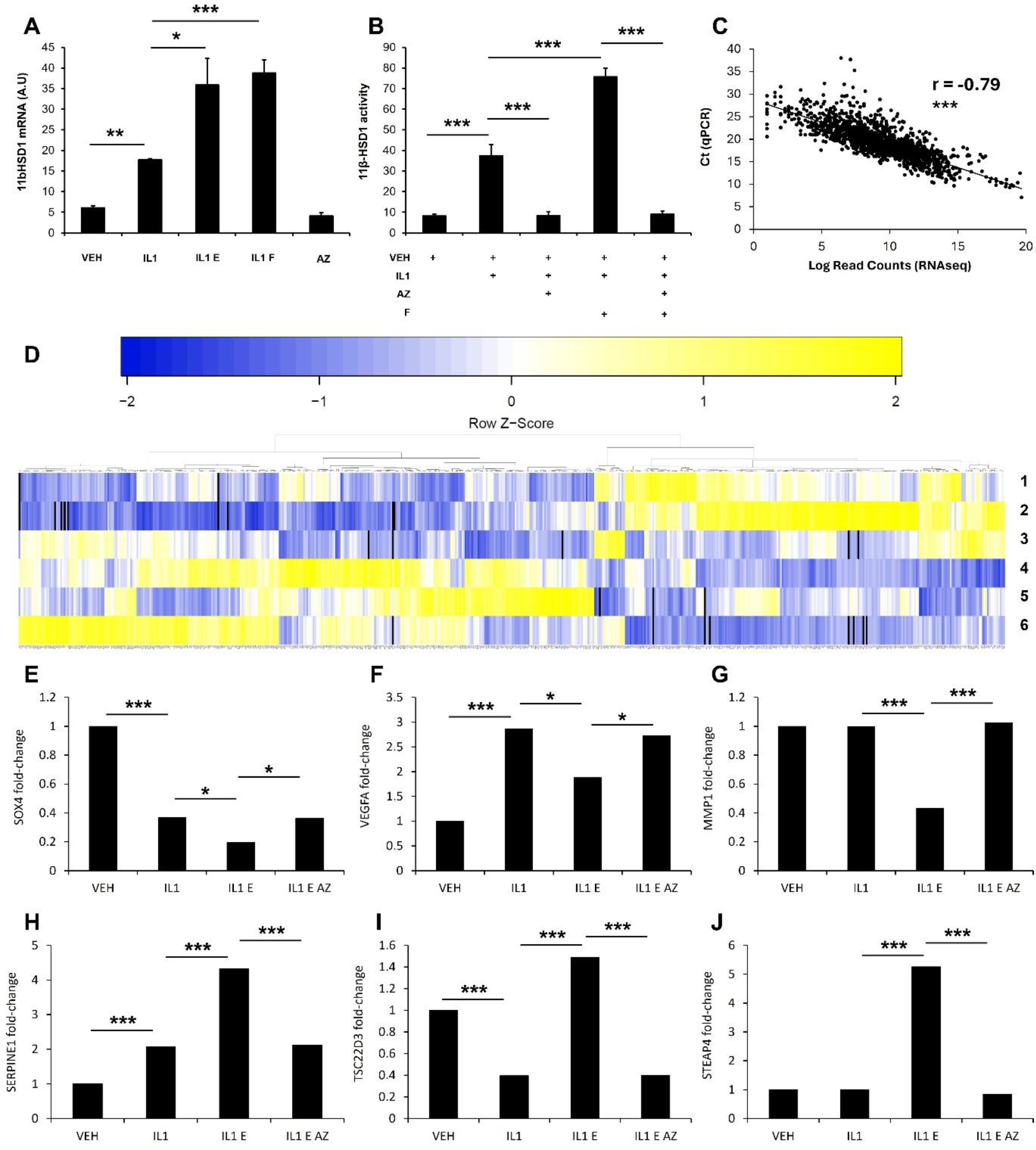
11β-HSD1 regulation, activity, and target genes in HDF **A)** 11β-HSD1 mRNA (by qPCR) following treatment with vehicle (VEH), IL-1β (IL1), IL-1β + cortisone (IL1 E), IL-1β + cortisol (IL1 F), or AZD4017 (AZ), n=3, **B)** 11β-HSD1 activity (% conversion of cortisone in 24 hours) following treatment with vehicle, IL-1β, IL-1β + AZD4017, IL-1β + cortisol, or IL-1β + cortisol + AZD4017, n=10, **C)** qPCR validation for 246 (of 611) 11β-HSD1 target genes and 32 control genes, representing 94.4% of the qPCR data, **D)** Heat map of 611 differentially expressed genes between HDF treated with IL-1β + cortisone (1-3) or IL-1β + cortisone + AZD4017 (4-6), **E-J)** Representative 11β-HSD1 target gene expression (by RNA-seq) demonstrating IL-1β antagonising, augmenting and independent effects (n=3). Significance * = p<0.05, ** = p<0.01, *** = p<0.001.

#### 3.1.2. 11β-HSD1 transcriptome

RNA-seq identified 611 gene targets for 11β-HSD1 in primary human dermal fibroblasts (Figure 1D, Supplemental Table 2). Of these, 357 were upregulated following 11β-HSD1 inhibition with AZD4017, including pro-inflammatory genes known to be suppressed by GC through 11β-HSD1 (IL1B, IL6, PTGS2, SOX4,TSC22D3), extracellular matrix proteoglycans (ITGA8, TNC), glycoprotein biosynthesis (GALNT12, SIGLEC15, ST3GAL1), collagen biosynthesis and remodeling (ADAMTS14, LOXL1, MMP1, MMP3, TIMP3), cell adhesion (ITGA1, ITGA2, ITGA3, ITGA7, ITGA8, ITGA11, LAMB3), and pro-angiogenic factors (VEGFA, VEGFC).

Conversely, 254 genes were downregulated by AZD4017, including known 11β-HSD1 target anti-inflammatory mediators (FKBP5, TSC22D3), EGF signaling (ANXA2, APPL2, PRKD1), blood clotting (F3, F10, PROS1, SERPINE1, SERPING1), protein glycosylation and pro-fibrotic regulators (ADAMTS2, ADAMTSL4, CCN2, CD163, GCNT1, TGFBR2, THSD4, ZEB1), and adipogenesis (IRS2, LEP, STEAP4).

11β-HSD1 target genes were either IL-1β antagonists (204 genes), IL-1β agonists (85 genes) or IL-1β independent (322 genes, Figure 1E-J, Supplemental Table 3). A further 1729 genes were found to be regulated by IL-1β only (Supplemental Table 3).

#### 3.1.3. 11β-HSD1 target genes

Over-representation analysis identified 386 pathways for genes upregulated by AZD4017 and 301 pathways for genes downregulated by AZD4017 (Supplemental Table 4). Most notable pathways included extracellular matrix organization (e.g., integrin cell surface interactions, focal adhesion, TGF-beta signaling, collagen biosynthesis and remodeling), inflammation (e.g., TNF and NF-kappa B signaling, IL-17 and IL-18 cytokines), angiogenesis (e.g., VEGF, plexin D1, nitric oxide, WNT, beta-catenin and NOTCH signaling, vascular endothelial cell function), hemostasis (e.g., fibrin clot formation, platelet adhesion and degranulation, complement and coagulation cascades), metabolism (e.g., IGF signaling, gastrin and bile acids, adipogenesis, lipolysis, insulin secretion, thermogenesis), and nerve fiber regulation (e.g., netrin-1 and ROBO signaling, axon guidance).

Results also indicated disease-specific pathways, particularly cardiovascular pathologies (e.g., cardiac arrhythmia and hypertrophy, cardiomyopathy, diabetic complications, atherosclerosis), extracellular matrix remodeling disorders (e.g., lung fibrosis, sclerosis), and inflammatory conditions (e.g., rheumatoid arthritis, viral infection).

### 3.2. Human skin

#### 3.2.1. Validation

45 11β-HSD1 target genes were common to both skin and HDF, displaying a positive correlation in regulation of gene expression by AZD4017 (Figure 2A, r = 0.47, p<0.01). 67 skin differentially expressed genes from the placebo group were also present in the HDF AZD4017 results, but as anticipated, their regulation did not correlate (Figure 2B, r = -0.2, p=0.10), validating our skin RNA-seq findings.

**Figure 2.**
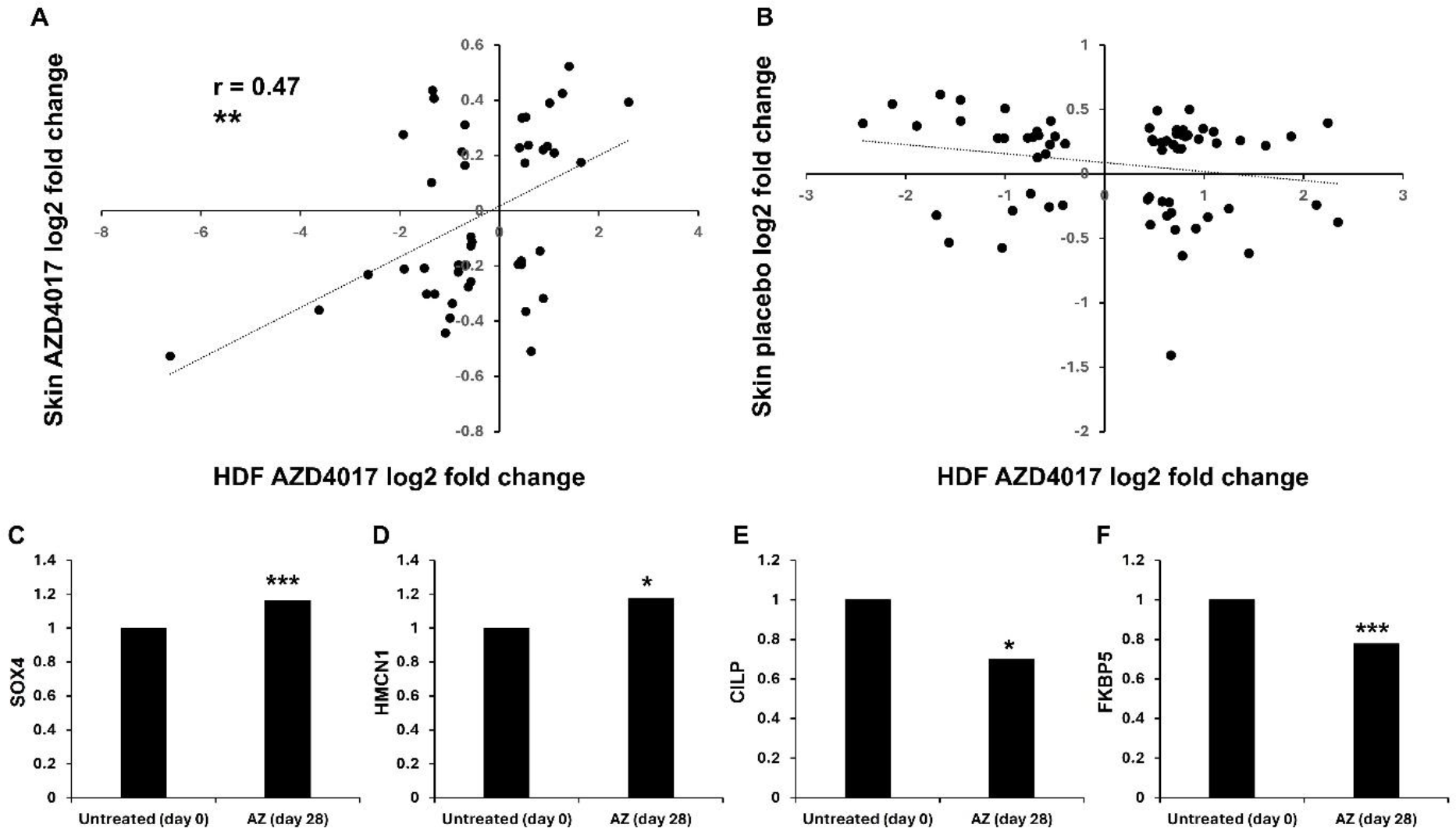
11β-HSD1 target genes in human skin **A)** Correlation between overlapping skin and HDF 11β-HSD1 target genes, n=45, **B)** Lack of correlation between skin from placebo-treated participants and overlapping HDF 11β-HSD1 target genes, n=67, **C-F)** Representative 11β-HSD1 target genes (by RNA-seq) in human skin, n=11 (omitting the five confounding participants). Significance * = p<0.05, ** = p<0.01, *** = p<0.001.

#### 3.2.2. 11β-HSD1 transcriptome

Differential expression analysis in human skin from people with type 2 diabetes treated with AZD4017 identified 1026 target genes (Figure 2C-F).

442 genes were upregulated by AZD4017, including pro-inflammatory (e.g., CX3CL1, GSDMD, IRF7, MAP4K4, SOX4, TNFRSF14), epidermal integrity (e.g., CLDN3, DSC2, DSG2, FGF10, HMCN1, KRT25, LPAR6, SLC12A8, SPHK2, SPRR2B), extracellular matrix composition (e.g., BAMBI, HAS3, MMP14, P3H3, PLOD1, THSD4, STAB2), epithelial cell differentiation (e.g., CITED4, EHF, KRT7), angiogenesis (e.g., APP, FLT1, PAN2, RUNX1, SEMA5A), coagulation (e.g., ADAMTS13, CEACAM1, DNASE1, ST3GAL4), lipid metabolism (e.g., AZGP1, CHKA, HMGCS2, PNPLA4, SMIM22, SREBF1), ion channels (e.g., ASIC1, CFTR, KCNA2, KCNA4, SGK1) and telomere maintenance (e.g., NEIL1, NOP10, TINF2).

Conversely, 397 genes were downregulated by AZD4017, including stress response (e.g., FKBP5, MEF2A), anti-inflammatory (e.g., ELF4, RC3H1, RPS6KA5, TSC22D3), IL-17 signaling (e.g., CALM1), epidermal barrier integrity (e.g., A2ML1, ALOXE3, CYP4F22, DSC1, DSG1, GRHL3, KRT5, KRT10, LGALS7, LGALS7B, LIPM, MAP2K3, MAPK13, TMPRSS11F, SCEL, SERPINB8), fibrosis (e.g., CILP, POSTN), lipid storage (e.g., APOB, ASAH2, CIDEC, DEGS2, GPAM, GPAT3, MEDAG), hypertension (e.g., CYP3A5), and blood clotting (e.g., F3, ITGB3, PRKCA).

A further 187 AZD4017 target genes were regulated in a comparable way to the placebo group and therefore omitted from subsequent pathway and correlation analyses. A total of 1136 genes were differentially expressed in the placebo group.

### 3.3. Human skin 11β-HSD1 target pathways

Over-representation analysis of upregulated genes in skin from people with type 2 diabetes treated with AZD4017 identified 73 pathways, whilst downregulated genes identified 411 pathways (Supplemental Table 5).

This included epidermal barrier (e.g., keratin and sialic acid metabolism, keratinocyte differentiation), cell adhesions (e.g., tight junctions), fibrosis (e.g., TGF-beta, AGE/RAGE signaling), inflammation (e.g., IL-7, IL-8, IL-17, MyD88, TNF and NF-kappa B signaling, transendothelial migration), glucose homeostasis (e.g., RAC1 signaling, insulin secretion, glycogen metabolism), lipid metabolism (e.g., sphingolipid, phospholipid, gastric acid secretion, adipocyte differentiation), angiogenesis (e.g., VEGF, ERBB, RUNX1 and WNT signaling, vascular permeability), blood pressure (e.g., angiotensin signaling, shear stress, endothelins), coagulation (e.g., hemostasis, platelet production, platelet calcium homeostasis, thrombin signaling), ion channels (e.g., calcium-activated potassium, TRP), and cell trafficking (e.g., kinesins). Comparable to our findings in dermal fibroblasts, disease-associated pathways regulated by 11β-HSD1 inhibition in human skin included cardiac hypertrophy and atherosclerosis.

### 3.4. 11β-HSD1 inhibitor efficacy

#### 3.4.1. 11β-HSD1 skin transcriptome

Of 814 11β-HSD1 skin target genes, 208 (25.6%) correlated (r >0.3 or <-0.3) with a change in systemic 11β-HSD1 activity (serum F/E) for AZD4017 and placebo-treated participants (Figure 3A-D). Correlations were significant for 46 genes (p<0.05), with a further 41 demonstrating a trend (p<0.1). Similar results were observed for cortisol levels (serum F/DHEAS, Figure 3E-H) and when combining 11β-HSD1 activity and cortisol levels (F/DHEAS*F/E, Figure 3I-L).

**Figure 3.**
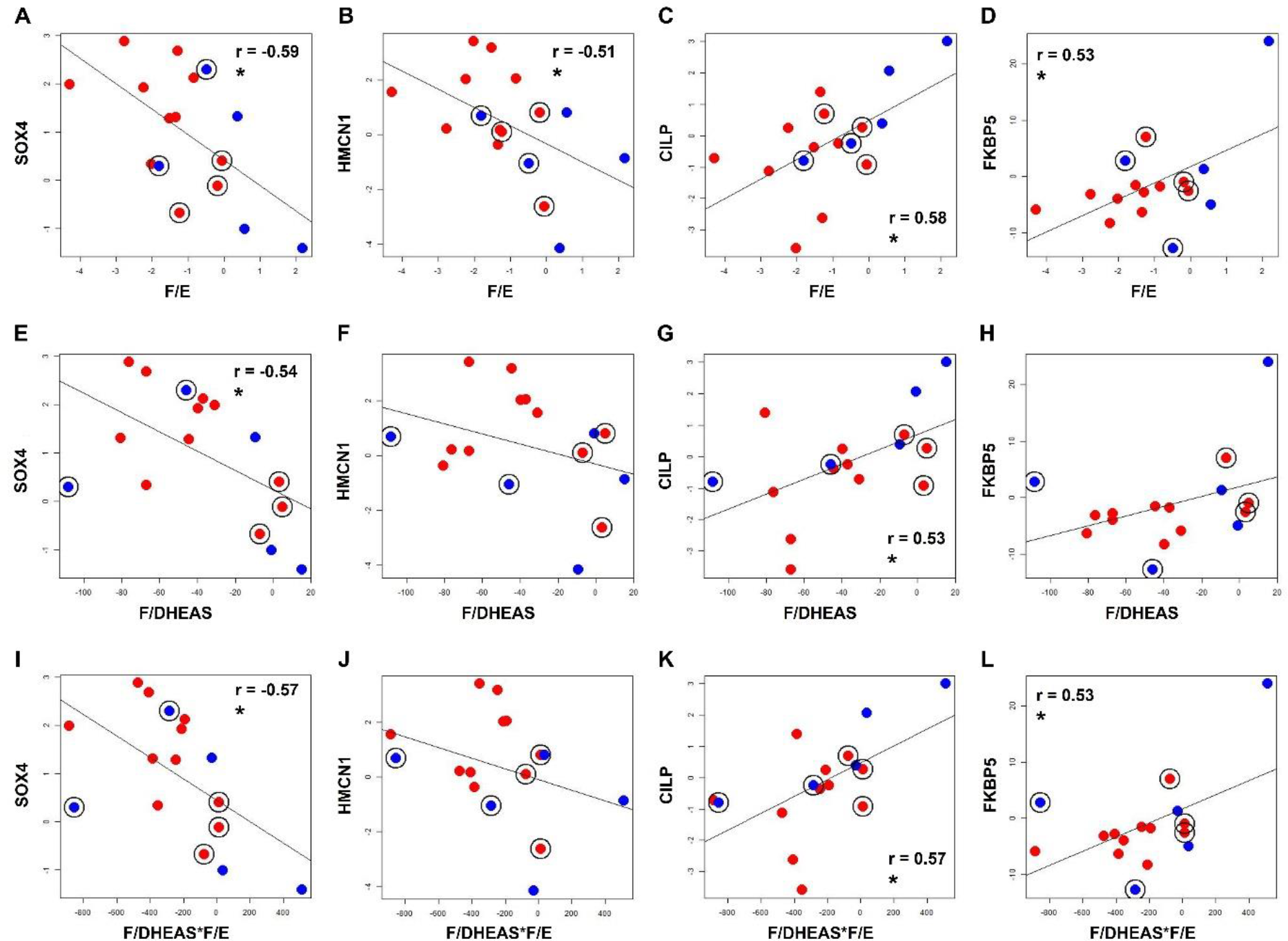
Regulation of 11β-HSD1 gene targets correlate with systemic cortisol and 11β- HSD1 activity Representative correlations (including the five confounding participants) between log2 fold- change in 11β-HSD1 target genes (by RNA-seq) and **A-D)** systemic 11β-HSD1 activity (serum F/E), **E-H)** systemic cortisol (serum F/DHEAS), and **I-L)** systemic 11β-HSD1 activity* systemic cortisol (serum F/E*F/DHEAS). Illustrates the confounding effect of three AZD4017-treated participants with relatively low baseline F/DHEAS (red with black outline) and two placebo-treated participants with a reduction in baseline F/DHEAS (blue with black outline). Red = AZD4017-treated, n=11, blue = placebo-treated, n=5. Significance * = p<0.05.

Notably, three AZD4017-treated participants with lowest baseline F/DHEAS cluster with placebo-treated participants, whilst two placebo-treated participants with a reduction in baseline F/DHEAS (as for AZD4107 treatment) cluster with AZD4017-treated participants (Figure 3). When these five participants are included in the RNA-seq analysis, AZD4017 efficacy for the top 20 most significantly upregulated and downregulated genes is reduced by 13.9±10% (p<0.001, n=23), demonstrating a significant confounder effect.

#### 3.4.1. Wound healing, blood pressure, and lipids

Similar findings were also observed for wound healing, which displayed a negative correlation with F/E (r = -0.65, p<0.01) and F/E*F/DHEAS (r = -0.61, p<0.01, Figure 4A-C). Percentage wound healing in AZD4017-treated participants was 1.48-fold higher than the placebo group (76.8±16.6 vs. 51.9±23.5, p<0.05), and increased by 10% to 1.63-fold (p<0.05) with the five confounders excluded.

**Figure 4.**
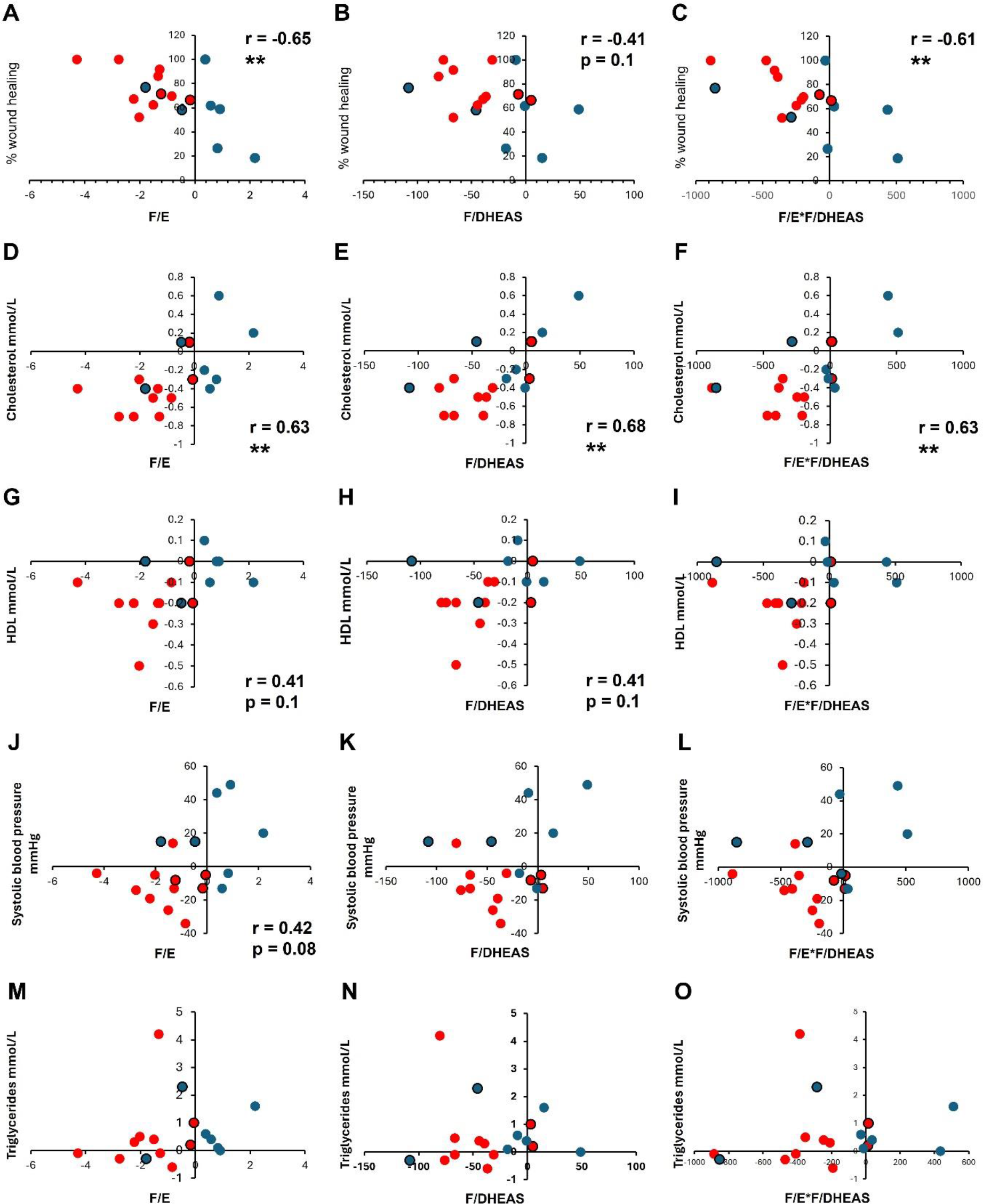
Regulation of wound healing, blood pressure and lipids by 11β-HSD1 correlates with systemic cortisol and 11β-HSD1 activity Correlations between systemic 11β-HSD1 activity (serum F/E), systemic cortisol (serum F/DHEAS), and systemic 11β-HSD1 activity* systemic cortisol (serum F/E*F/DHEAS) for **A-C)** wound healing (AZD4017 n=10, placebo n=7), **D-F)** cholesterol (AZD4017 n=10, placebo=7), **G-I)** high density lipoprotein (AZD4017 n=9, placebo n=7), **J-L)** systolic blood pressure (AZD4017 n=11, placebo n=7), **M-O)** triglycerides (AZD4017 n=10, placebo n=7). As for 11β-HSD1 transcriptomic targets, the same confounding effect of three AZD4017- treated participants (red with black outline) and two placebo-treated participants (blue with black outline) is observed. Red = AZD4017-treated, blue = placebo-treated. Significance ** = p<0.01.

Conversely, positive correlations (and trends) were found for changes in cholesterol (F/E; r = 0.63, p<0.01, F/DHEAS; r = 0.68, p<0.01, F/E*F/DHEAS; r = 0.63, p<0.01, Figure 4D-F), high density lipoprotein (F/E; r = 0.41, p = 0.1, F/DHEAS; r = 0.41, p = 0.1, Figure 4G-I), and systolic blood pressure (F/E; r = 0.42, p = 0.08, Figure 4J-L). AZD4017 reduced cholesterol by -0.47 ± 0.2 mmol/L compared to placebo (-0.02 ± 0.3, p<0.001), with similar reduction for high density lipoprotein (-0.19 ± 0.12 mmol/L vs. -0.04 ± 0.09, p<0.01) and systolic blood pressure (-10.7 ± 11.9 mmHg vs. 7.6 ± 21.7, p<0.05). These effects increased by 14.9% (-0.54, p<0.001), 9.5% (-0.21, p<0.01), and 5.6% (-11.3, p = 0.052), respectively, with the five confounders excluded. However, triglycerides were not regulated by AZD4017 (0.39 ± 1.38 mmol/L vs. 0.44 ± 0.57, p=0.91, Figure 4M-O) and were unaffected by confounder exclusion.

## 4. DISCUSSION

Improving our understanding of 11β-HSD1 molecular targets and therapeutic efficacy is vital for identifying new clinical indications and developing more targeted treatment strategies.

Here, we used the selective 11β-HSD1 inhibitor AZD4017 for the first human transcriptomic profiling of 11β-HSD1 targets. Transcriptomic targets confirmed serum cortisol as an important biomarker of AZD4017 responsiveness, further corroborated by functional wound healing and cardiometabolic outcomes in people with type 2 diabetes. Our findings also shed new light on serum cortisol as a biomarker for 11β-HSD1 inhibitor efficacy for more robust AZD4017 targeting in future clinical studies.

GC delay wound healing through multiple mechanisms spanning all stages of the repair process; haemostasis, inflammation, proliferation (re-epithelialization), and extracellular matrix remodelling[18, 19]. Here, we demonstrate that 11β-HSD1 inhibition displays a pro-healing transcriptional profile, supporting our recent clinical findings[7, 20] and therapeutic potential in skin[21]. Notably, genes associated with healing in diabetic foot ulcers such as downregulation of CD163 and upregulation of MMP1, MMP3, CHI3L1, TNFAIP6, TCF7, IL1B, IL6, CCL2, CXCL12, PTGS2, VEGFA[22] were regulated in the same manner by AZD4017, highlighting the importance of inflammation and limiting fibrosis to effective wound healing[23].

GC are well-known for their anti-inflammatory effects, supporting our observed induction of pro-inflammatory genes and pathways by AZD4017. Importantly, this has not translated to adverse outcomes during mild / acute inflammation[7, 11, 24], but could be disadvantageous during severe / chronic inflammation[25, 26].

Interestingly, AZD4017 also suppressed pro-inflammatory genes including PTGDR (mast cell degranulation e.g., asthma), FOXO3 (regulatory T-cell differentiation), CXCL5 (neutrophil activation), MMD (macrophage maturation), and SOX4 (IL-17 promotor), supported by emerging evidence that GC promote inflammation in certain contexts[27].

Therefore, the role of 11β-HSD1 in conditions exacerbated by psychological stress such as atopic dermatitis and eczema[28] also warrants investigation, particularly as a significant number of skin barrier (epidermal) genes were also regulated, supporting our previous findings that GC promote barrier function[7, 11].

GC excess also drives many clinical features of cardiometabolic morbidity and mortality[1, 29] but knowledge on how 11β-HSD1 regulates these processes remains sparse. Alongside identifying novel 11β-HSD1 transcriptional targets relevant to improved wound healing and skin function, our findings provide new mechanistic evidence for wider therapeutic benefit. For example, lower expression of pro-fibrotic POSTN and CILP (reduced by AZD4107 here) were associated with improved cardiac remodelling and function[30, 31], whilst 11β-HSD1 inhibition also improved recovery following myocardial infarction in mice [31, 32]. Further, we found that AZD4017 induced thromboprotective ADAMTS13, PROCR, STAB2, PLAU, and reduced prothrombotic SERPINE1, with differential expression of other coagulation genes (PROS1, PLAT, VWF, ST3GAL4, RUNX1) also suggesting a novel role for 11β- HSD1 in the regulation of haemostasis[33–35]. Other novel 11β-HSD1 targets include AZD4017-upregulated SOSTDC1; a TGF-beta inhibitor associated with endometrial receptivity[36], PIEZO2; a cation channel associated with mechanosensation and diabetic neuropathy[37, 38], and key regulators of metabolism such as GPAT3[39, 40], STEAP4[41], HMGCS2, and SOX4[42].

Previous selective 11β-HSD1 inhibitor clinical trials found modest improvements in HbA1c[3, 4], fasting plasma glucose[3], HOMA-IR[3], body weight[3–6], blood pressure[4, 7], lipids[3, 5, 7, 8], hepatic steatosis[9, 10], lean muscle mass[8] and wound healing[7], but failed to progress to phase 3 trials due to limited efficacy.

Our findings suggest that baseline cortisol levels (serum F/DHEAS) and 11β-HSD1 activity (serum F/E) may predict 11β-HSD1 inhibitor efficacy, such that people with higher cortisol levels would benefit more from 11β-HSD1 inhibition. Further, we demonstrate how changes in cortisol levels in the placebo group can also skew 11β-HSD1 efficacy. The lack of adjustment for these important variables may explain the relatively modest effect sizes in previous 11β-HSD1 inhibitor studies. However, one study reported a correlation between change in serum F/E and change in lumbar puncture pressure (primary outcome) in the 11β- HSD1 inhibitor arm[15], supporting our findings. This is also in agreement with greater 11β- HSD1 inhibitor efficacy in studies with more consistent GC excess, such as Cushing’s disease or exogenous GC therapy[24, 43].

Our findings have implications beyond wound healing, and evidence is growing for cortisol as a driver of disease severity in diabetes[44, 45]. In the context of our 11β-HSD1 transcriptomic profile, F/DHEAS and F/E may be useful indices in assessing disease progression in other settings, such as cardiovascular morbidity which is strongly associated with GC excess[29]. Extended dosing studies would be required, as 11β-HSD1 inhibitors are currently limited to a maximum dosing duration of 3 months. As 11β-HSD1 is elevated in older people[46–48], these indices may also be useful in predicting age-associated health outcomes.

Limitations of our work include a small sample size (n=14 per arm); larger confirmatory studies in people with healing and non-healing diabetic foot ulcers are now required (for safety, our initial clinical study focused on acute wound healing). The application of spatial omics to refine the 11β-HSD1 transcriptomic profile *in situ* would also be of great interest, as our study was limited to bulk skin expression – this also likely explains the relatively modest gene regulation and associated correlations.

In summary, this study presents new mechanistic detail of 11β-HSD1 target genes relevant to wound healing alongside wider therapeutic applications. It also presents a more robust way to evaluate 11β-HSD1 inhibitor efficacy by taking the degree of GC excess into consideration, paving the way for improved therapeutic targeting in future clinical studies.

## ACKNOWLEDGEMENTS AND FUNDING

This work was funded by a Medical Research Council Confidence in Concept Award to AT (MC_PC_15046), NIHR Senior Investigator Award to PMS (NF-SI-0514-10090), and the NIHR Leeds Biomedical Research Centre. Study registration at https://doi.org/10.1186/ISRCTN74621291 (ISRCTN74621291 18/10/2017) and https://clinicaltrials.gov/ct2/show/NCT03313297 (NCT03313297 18/10/2017).

## 6. DATA AVAILABILITY

The authors confirm that the data supporting the findings of this study are available within the article and its supplementary materials.

---

**Supplemental Figure 1:**
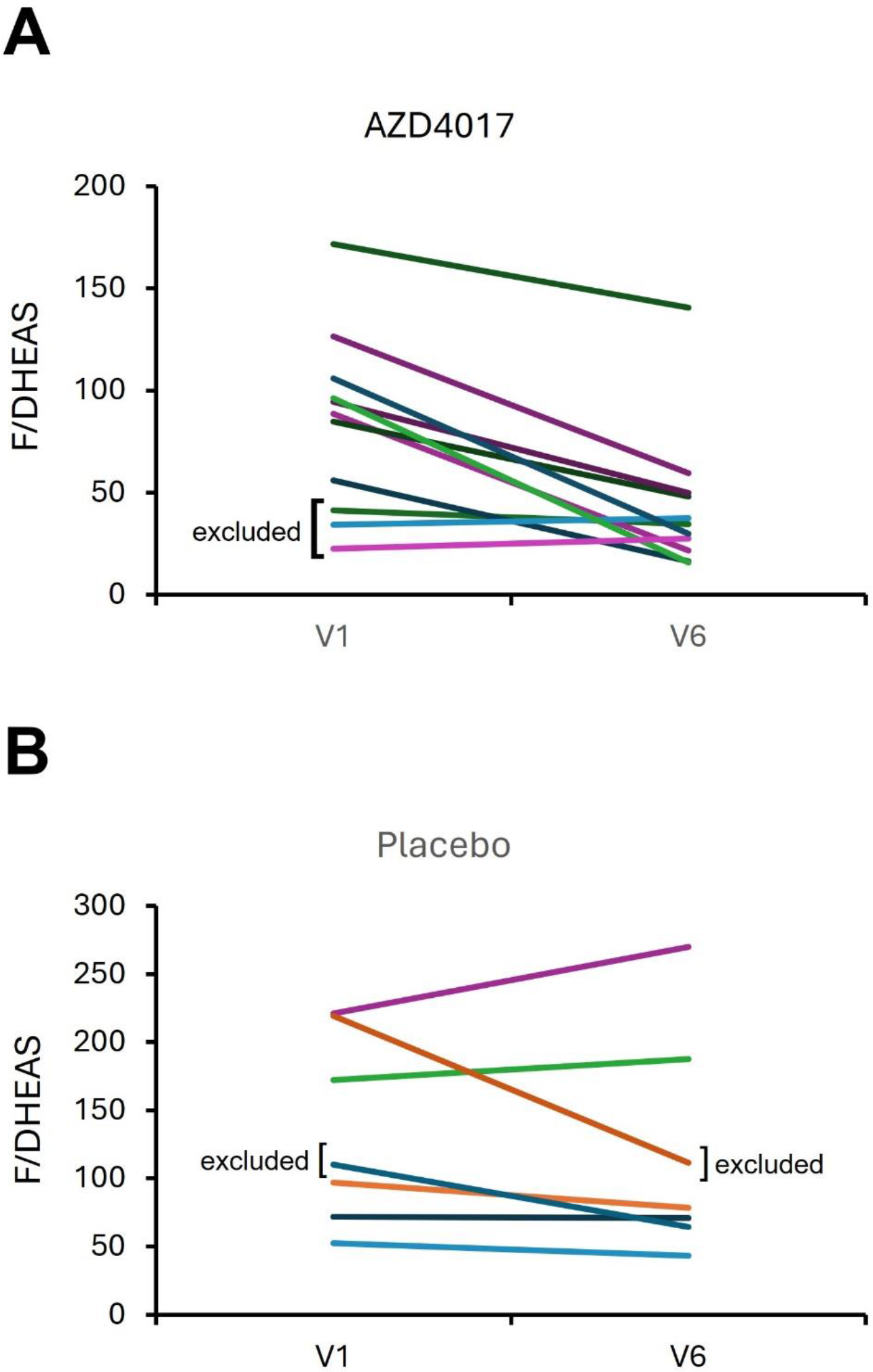
Change in serum cortisol (F) and dehydroepiandrosterone sulfate (DHEAS) in people treated with AZD4017 **(A)** or placebo **(B).** Illustrates five participants excluded from the initial transcriptomic analysis but included in the subsequent llβ-HSDl target gene correlations with changing cortisol levels.

